# Prognostic Significance of Different Ventricular Ectopic Burdens During Exercise in Asymptomatic UK Biobank Subjects

**DOI:** 10.1101/2023.03.04.23286713

**Authors:** Stefan van Duijvenboden, Julia Ramírez, Michele Orini, Aiden Doherty, Andrew Tinker, Patricia B. Munroe, Pier D. Lambiase

## Abstract

**Background and Aims:** The consequences of exercise-induced premature ventricular contractions (PVCs) in asymptomatic individuals remain unclear. This study aimed to assess the association between PVC burdens during stress testing and major adverse cardiovascular events (MACE; myocardial infarction (MI), life-threatening ventricular arrhythmia (LTVA), and heart failure (HF)), and all-cause mortality. Additional end-points were: MI, LTVA, HF and cardiovascular mortality.

**Methods:** A neural network was developed to count PVCs from ECGs recorded during exercise (6 min) and recovery (1 min) in 48,502 asymptomatic participants from UK Biobank. Associations were estimated using multivariable Cox proportional hazard models.

**Results:** Mean age was 56.8 (+/-8.2 years); 51.1% were female, and median follow-up was 11.5 years. Low PVC counts during exercise and recovery were both associated with MACE risk, independently from clinical factors: adjusted hazard ratio [HR]: 1.2 (2-4 exercise PVCs, p = 0.022) and HR 1.2, (1 recovery PVC, p = 0.031). Risk increased with increasing PVC count: HR 1.8 (>25 exercise PVCs, p<0.001) and HR 1.5 (≥5 recovery PVCs, p < 0.001). A similar trend was observed for all-cause mortality, although associations were only significant for higher PVC burdens: HRs: 1.4 (11-25 exercise PVCs, p = 0.007) and 1.5 (≥5 recovery PVCs, p < 0.001). Complex PVCs rhythms were associated with higher risk compared to PVC count alone. PVCs were also strongly associated with incident HF, LTVA, and cardiovascular mortality, but not MI.

**Conclusion:** PVC count during exercise and recovery are both associated with MACE, all-cause mortality, HF, LTVAs and cardiovascular mortality, independent of clinical and exercise stress test factors, indicating a “dose response” between PVC count and risk. Complex PVCs rhythms are associated with higher risk compared to PVC count alone.

## Introduction

Exercise stress testing poses a dynamic physiological stress on the heart that may unmask underlying cardiac anomalies not evident at rest and is commonly ordered to guide risk assessment in low or intermediate risk individuals^1^. Premature ventricular complexes (PVCs) are commonly observed during exercise stress testing (prevalence ∼7%)^2^. The prognostic implications of high PVC burden are well-recognised in patients with structural heart disease ^3–7^, however the implications for asymptomatic individuals remain incompletely understood. As highlighted recently, the increased availability of wearable ECG monitoring devices has significantly improved the identification of asymptomatic subjects with PVCs, emphasising the need to better understand the association between PVCs and cardiovascular risk^8^.

The prognostic implications of low and intermediate PVC counts are unknown, and little is known about the specific risks carried by different PVCs rhythms (e.g. couplets, triplets, and bigeminy). A very limited number of studies have investigated the prognostic implications of PVCs on exercise in asymptomatic individuals ^8–14^. The latest evidence suggests that high-grade PVCs (frequent or complex PVCs) during *recovery after exercise* are associated with a 1.7x increased long-term cardiovascular mortality risk, independent of established clinical risk factors, whereas no significant association was found for PVCs *during* exercise in 5,486 individuals of which 311 died due to cardiovascular disease^8^. However, the quality of the evidence is limited because study cohorts are small, usually not population based, and, as highlighted previously, there is lack of uniformity among definitions used to designate frequent PVCs ^15^. In addition, little is known about the association of PVCs with other important cardiac outcomes including myocardial infarction, life threatening ventricular arrhythmia (LTVA), and heart failure (HF). We hypothesised that the risk associated with PVCs increases with frequency of PVCs and their pattern.

Therefore, we investigated exercise induced PVCs in 48,502 asymptomatic individuals without known cardiovascular disease with over 10 years follow-up from the UK Biobank (UKB) study to address the following hypotheses: 1. PVC burden during exercise (N= 1,2-4,5-10,11-25,>25 beats) and recovery (N = 1,2-4,5) determine the clinical adjusted risk for major adverse cardiovascular events (MACE) and all-cause mortality. 2. PVC burden is associated with myocardial infarction, LTVA, and heart failure (HF), and cardiovascular mortality. 3. Levels of risk differ according to the different PVC patterns.

## Methods

### UK Biobank

The UK Biobank is a large population cohort established between 2006 and 2010, with even numbers of men and women aged 40-69 years on recruitment from 21 assessment centres across England, Wales, and Scotland. The study has extensive baseline and follow-up clinical, biochemical, and outcome measures and approval from the North West Multi-Centre Research Ethics Committee. All participants provided informed consent at the time of enrolment for their data to be linked to the health record systems.

### Exercise stress test

From the full UK Biobank cohort, 95,154 participants consented to participate in the exercise test between 2009 - 2013. The test used cycle ergometry on a stationary bike (eBike, Firmware v1.7) in conjunction with a single lead (lead I) electrocardiograph device (CAM-USB 6.5, Cardiosoft v6.51, sample rate 500Hz). The exercise protocol consisted of a 15s pre-test resting phase, followed by graded activity (6 min) and a recovery period with hands remaining on the handlebars whilst remaining still and silent (1 min, no cool-down period). According to protocol, participants were risk-stratified into groups and those who were allowed to exercise were categorized to perform a test at either 35% or 50% of the maximum predicted workload depending on their risk profile (https://biobank.ndph.ox.ac.uk/showcase/ukb/docs/Cardio.pdf). The predicted absolute maximum workload was calculated according to a formula, which includes age, height, weight, resting heart rate and sex.

### ECG analysis

As we did not had access to a method to detect and count premature ventricular contractions (PVCs), we developed, trained and validated a deep-learning model to detect PVCs from raw single-lead ECG recordings from the exercise cohort in the UK Biobank study. ECGs were first bandpass filtered (0.5 – 45Hz), down sampled to 250Hz and then split in non-overlapping segments of 2048 time samples (∼8.2s) for computational efficiency. A 1D convolutional neural network (CNN) was trained on the ECG segments to estimate the probabilities for each time sample in the segment that it belonged to (i) a normal (narrow) QRS complex, (ii) a PVC QRS morphology, and (iii) neither of both (Central illustration). The network was implemented using the Keras framework with a Tensorflow (Google) backend and Python (v3.8.5) ^16^. The main architecture was composed of two 1D convolutional layers connected to a drop out, dense and classification layer. Post processing was applied to re-join segments and to filter the estimated probabilities to improve localisation and classification of QRS complexes in the ECG.

The network was trained on 41,025 ECG segments from 723 participants who had at least 1 PVC during the exercise test identified following visual inspection of exercise ECGs from 1142 participants with abnormal RR intervals from data previously derived by our group ^17^. The performance algorithm was tested in a dataset of 79,257 ECG segments from 1500 participants that were randomly selected from the remaining ECGs in UK Biobank from participants without cardiovascular disease (Figure 1). All ECGs in training and testing sets were manually reviewed by an expert (SVD) to localise all QRS complexes and to label them as normal beats or PVC. A PVC was defined as a QRST complex with QRS duration ≥0.12 seconds not preceded by at least 0.12 seconds by a P wave.

**Figure 1:**
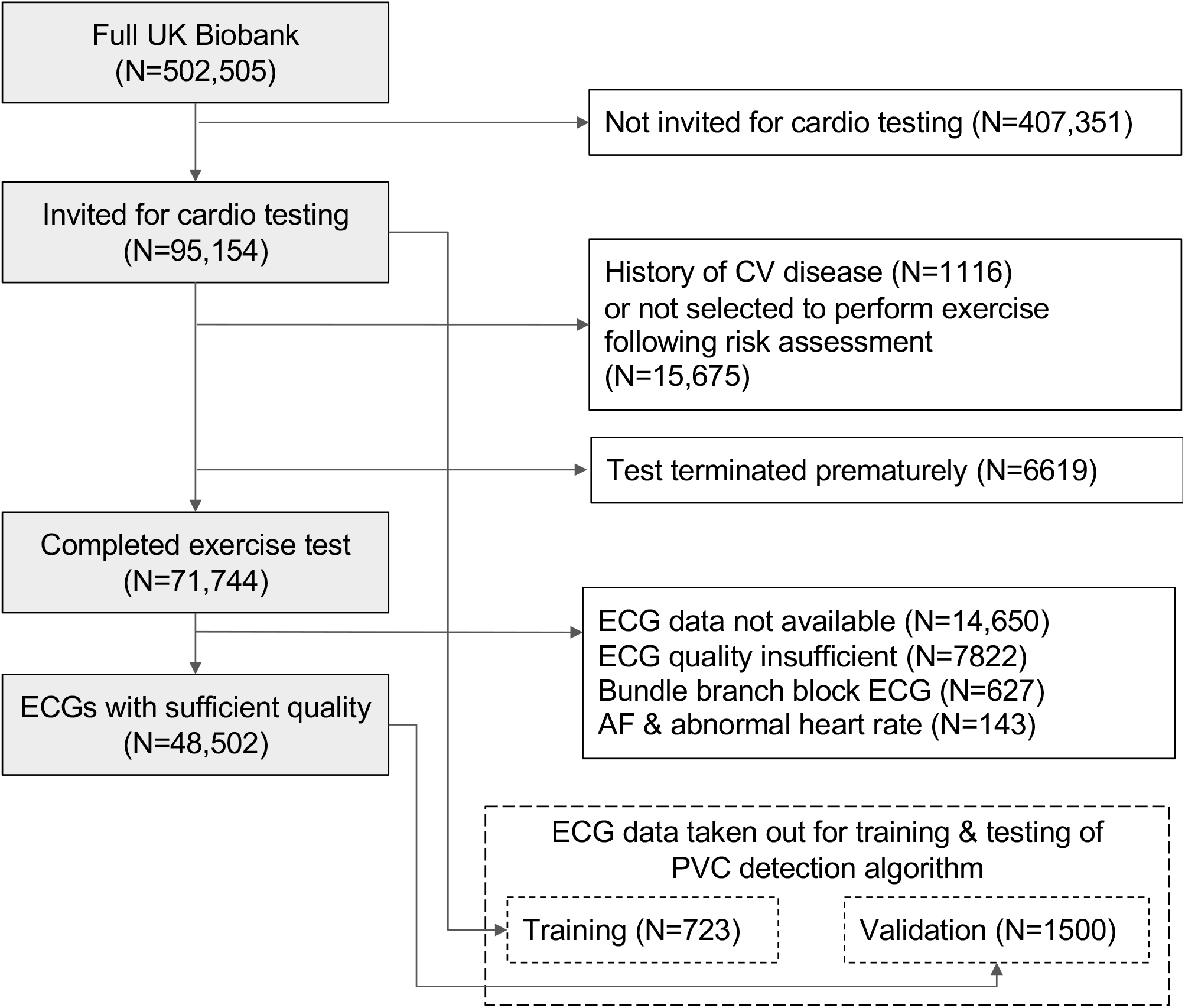
Study exclusion diagram. History of cardiovascular (CV) disease was identified using self-reported and hospital episode data (Supplemental Table 1). ECG = electrocardiogram, AF = atrial fibrillation.

Importantly, the distribution of PVC count was highly skewed (Supplemental Figure 1) with excessive zero PVC counts in most participants (xx% having 0 PVCs). We therefore evaluated the algorithm’s performance to accurately predict 6 pre-defined categories (0, 1, 2-4, 5-10, 11-25, and >25 PVCs), which were chosen based on clinical utility. Satisfactory performance was achieved: in the validation dataset, the overall accuracy was 98.5%, and sensitivity and positive predictive values ranged between 91.4 – 99.0% and 92.6 – 99.8%, respectively, across the PVC count categories (Supplemental Table 1). The algorithm was used to analyse all remaining ECGs in our dataset that were not part of training or validation set.

To compare our findings with the most recently published work from Refaat et al, we reviewed ECGs with PVCs for high-grade PVCs using the same criteria: frequent (>10 per minute), multifocal, >2 consecutive PVCs in a row (including ventricular tachycardia), or R-on-T type ^8^. R-onT and multifocal PVCs were manually confirmed by reviewing all ECGs with short PVC coupling interval (<400 ms) or low correlation among PVC beats (correlation coefficient <0.75).

### Ascertainment of study endpoints

All UKB participants consented to be followed-up through linkage to their health-related records and the UK death register ^18^. Participants with known cardiovascular disease prior to the exercise stress test, identified using either health-recorded or self-reported in the baseline questionnaire, were excluded (codes are provided in Supplemental Table 2). The primary study endpoint was major adverse cardiovascular events (MACE), defined as either hospitalisation or death due to myocardial infarction, HF, or LTVA. Cases were identified using relevant International Classification of Diseases 10th Revision (ICD10) or Operating procedures (OPSC4) codes in the health-related records or death register (Supplemental Table 3). The secondary endpoint was all-cause mortality. We also performed two additional analyses: 1) to evaluate the associations with myocardial infarction, LTVA, and HF separately, and 2) the association with cardiovascular death. The follow-up period was determined by the first appearance of ICD10 or OPSC4 codes in either heath record or death register data. Participants who did not experience an event were censored at death or the end of the follow-up period (November 12^th^, 2021).

### Statistical analysis

Descriptive statistics are presented as mean ± standard deviation (SD) for continuous variables or frequency (percentage) for categorical variables. Baseline clinical and exercise ECG characteristics were compared between each of the group with more than 1 PVC (1, 2-4, 5-10, 11-25, and >25 PVCs, for exercise, and 1, 2-4, ≥5 for recovery) and the group with no PVCs. Bonferroni correction was applied for multiple testing (n=5 for exercise PVCs and n=3 for recovery PVCs). The PVC counts had been pre-selected based on clinical utility. We used Wilcoxon rank-sum tests for continuous variables and the chi-square or Fisher exact test for categorical variables, whichever was appropriate. We constructed survival probability curves for MACE and all-cause mortality stratified for PVC count. The association between PVC count and study endpoints was investigated using multivariable-adjusted Cox proportional hazards regression. A minimally adjusted analysis used sex, age, and the number of heart beats during exercise or recovery. In a second model we addressed potential sources of confounding by further adjusting for clinical variables (hypertension, type 2 diabetes, LDL and HDL cholesterol, triglycerides, smoking, body mass index) and (exercise) ECG variables (QTc interval, QRS duration, ST-segment depression during exercise of >0.1mV, and heart rate increase during exercise (for exercise PVCs) or decrease during recovery (for recovery PVCs). Hypertension was defined as systolic blood pressure of ≥140 mmHg or diastolic blood pressure of ≥90 mmHg measured on the day of assessment ^19^ or a previous a diagnosis of hypertension (details provided in Supplemental Table 4). Inspection of Schoenfeld residuals revealed a possible violation of the proportional hazards assumption by sex. To avoid potential bias, we stratified Cox models by sex.

Missing variables (smoking status (0.4%), low- and high-density lipoprotein (LDL, HDL) cholesterol (8.2 and 13.2%, respectively), triglycerides (8.2%), BMI (<0.01%), QRS duration (0.9%), QTc interval (4.6%), ST depression (0.02%)) were imputed using the multiple imputation by chained equations approach, with five imputed datasets and ten iterations ^20^. For each variable we specified a predictive mean matching model, and we used all baseline variables to inform the imputation. Imputations were found acceptable by comparison of plots of the distribution of recorded and imputed values for all measurements. Cox regression models were estimated separately for each imputed dataset and then pooled together to obtain one overall set of estimates. A *P* < 0.05 was considered statistically significant. Statistical analyses were performed in R v4.0.2 ^21^.

### Sensitivity analyses – confounding effects of high PVC burden at rest

We performed sensitivity analyses to investigate whether the observed associations between exercise induced PVCs and MACE and all-cause mortality were independent from high PVC burden at rest. In this analysis, we added a binary covariate to the fully-adjusted cox models, indicating whether the participant had high PVC burden at rest, defined as having ≥1 PVC recorded during the available 15s (pre-exercise) resting period.

## Results

Following exclusions for reasons highlighted in Figure 1, 48,502 individuals without known cardiovascular disease and with complete ECGs were included in the study. The study population comprised a balanced set of middle-aged men and women (51.1% female, mean age 56.8 (+/- 8.2) years). Table 1 summarizes the number of participants in each PVC count category during exercise (A) and recovery (B). Participants with higher PVC counts were older, more likely to be male, diabetic, and hypertensive. They were also more likely to show ST-segment depression (>0.1mV). After a median follow-up period of 11.5 years (IQR: 3.3 months), there were 1960 (4.0%) MACE events (hospitalisation or death) and 2067 (4.3%) participants died from any cause.

**Table 1A:**
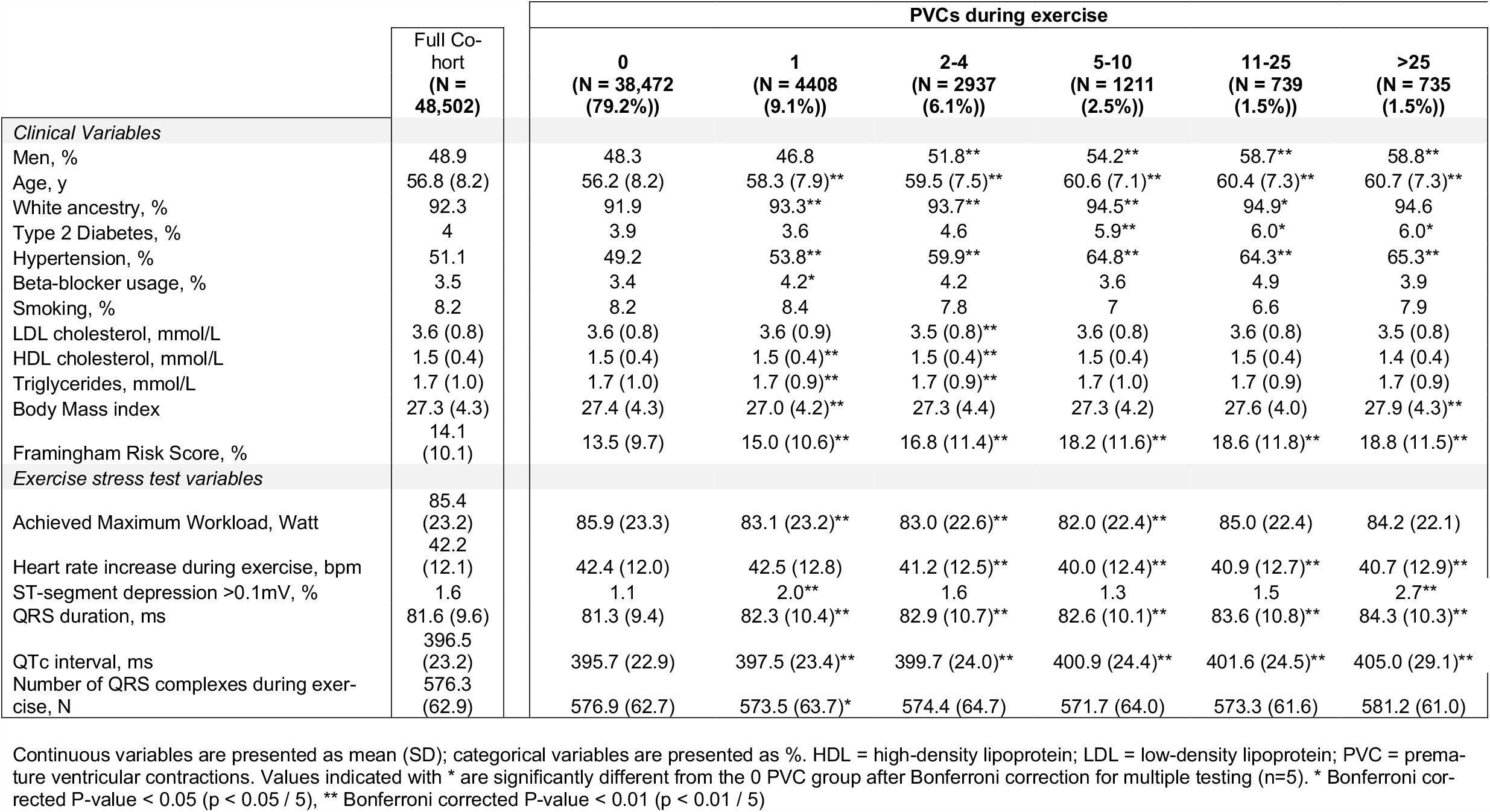
Baseline Clinical and Stress Test Characteristics During Exercise

**Table 1B:** Baseline Clinical and Stress Test Characteristics During Recovery

|  | Full Cohort<br>(N = 48,502) | PVCs during Recovery |  |  |  |
| --- | --- | --- | --- | --- | --- |
|  |  | 0<br>(N = 43,655 (90.0%)) | 1<br>(N = 2563 (5.3%)) | 2-4<br>(N = 1397 (2.9%)) | ≥5<br>(N = 887 (1.8%)) |
| *Clinical Variables* |  |  |  |  |  |
| Men, % | 48.9 | 48.7 | 49.3 | 51.6 | 52 |
| Age, y | 56.8 (8.2) | 56.5 (8.2) | 58.8 (7.8)** | 60.1 (7.4)** | 60.3 (7.6)** |
| White ancestry, % | 92.3 | 92.1 | 94.5** | 94.5** | 94.7* |
| Type 2 Diabetes, % | 4 | 3.9 | 5.2** | 5.3* | 4.7 |
| Hypertension, % | 51.1 | 50.2 | 55.5** | 62.8** | 63.6** |
| Beta-blocker usage, % | 3.5 | 3.5 | 3.6 | 3.6 | 3.6 |
| Smoking, % | 8.2 | 8.2 | 8.3 | 6.7 | 7.5 |
| LDL cholesterol, mmol/L | 3.6 (0.8) | 3.6 (0.8) | 3.6 (0.8) | 3.6 (0.8) | 3.6 (0.8) |
| HDL cholesterol, mmol/L | 1.5 (0.4) | 1.5 (0.4) | 1.5 (0.4)** | 1.5 (0.4)* | 1.5 (0.4) |
| Triglycerides, mmol/L | 1.7 (1.0) | 1.7 (1.0) | 1.7 (0.9) | 1.7 (1.0) | 1.7 (0.9) |
| Body Mass index | 27.3 (4.3) | 27.4 (4.3) | 27.1 (4.3)** | 27.3 (4.2) | 27.3 (4.1) |
| Framingham Risk Score, % | 14.1 (10.1) | 13.9 (10.0) | 16.0 (11.4)** | 17.3 (11.6)** | 17.5 (10.7)** |
| *Exercise stress test variables* |  |  |  |  |  |
| Achieved Maximum Workload, Watt | 85.4 (23.2) | 85.6 (23.2) | 83.3 (22.8)** | 83.0 (22.0)** | 82.5 (22.6)** |
| Heart rate recovery, bpm | 29.5 (10.1) | 29.7 (10.1) | 28.2 (10.4)** | 28.5 (10.8)** | 26.9 (11.7)** |
| ST-segment depression >0.1mV, % | 1.3 | 1.2 | 1.5 | 1.8 | 2.6** |
| QRS duration, ms | 81.6 (9.6) | 81.4 (9.5) | 82.3 (10.4)** | 83.4 (10.8)** | 83.9 (10.8)** |
| QTc interval, ms | 396.5 (23.2) | 396.0 (23.0) | 399.0 (23.5)** | 402.3 (24.6)** | 404.2 (28.2)** |
| Number of QRS complexes during recovery, N | 89.3 (13.1) | 89.2 (13.2) | 89.7 (13.2) | 88.9 (12.7) | 90.7 (12.5)** |

Continuous variables are presented as mean (SD); categorical variables are presented as %. HDL = high-density lipoprotein; LDL = low-density lipoprotein; PVC = premature ventricular contractions. Values indicated with \* are significantly different from the 0 PVC group after Bonferroni correction for multiple testing (n=5). \* Bonferroni corrected P-value < 0.05 (p < 0.05 / 5), \*\* Bonferroni corrected P-value < 0.01 (p < 0.01 / 5)

### PVCs during exercise

The incidence of MACE events and all-cause mortality were significantly higher in participants with PVCs during exercise and increased with PVC count. The incidence for MACE for individuals without PVCs was 3.6% compared to 4.6% in the group with 1 PVC (p = 0.001). Among participants with PVCs, the incidence further increased to 9.4% (> 25 PVCs) and was significantly higher compared to the group with 1 PVC (p < 0.001). A similar trend was observed for all-cause mortality: the incidence increased from 3.9% (0 PVCs) to 4.7% (1 PVC, p = 0.009). Among participants with PVCs, the incidence increased from 4.7% (1 PVC) to 9.1% (> 25 PVCs, p < 0.001). The unadjusted Kaplan-Meier curves showed decreasing survival rates for both MACE and all-cause mortality with increasing PVC counts (p < 0.001, Figure 2, Supplemental Figure 2)).

**Figure 2:**
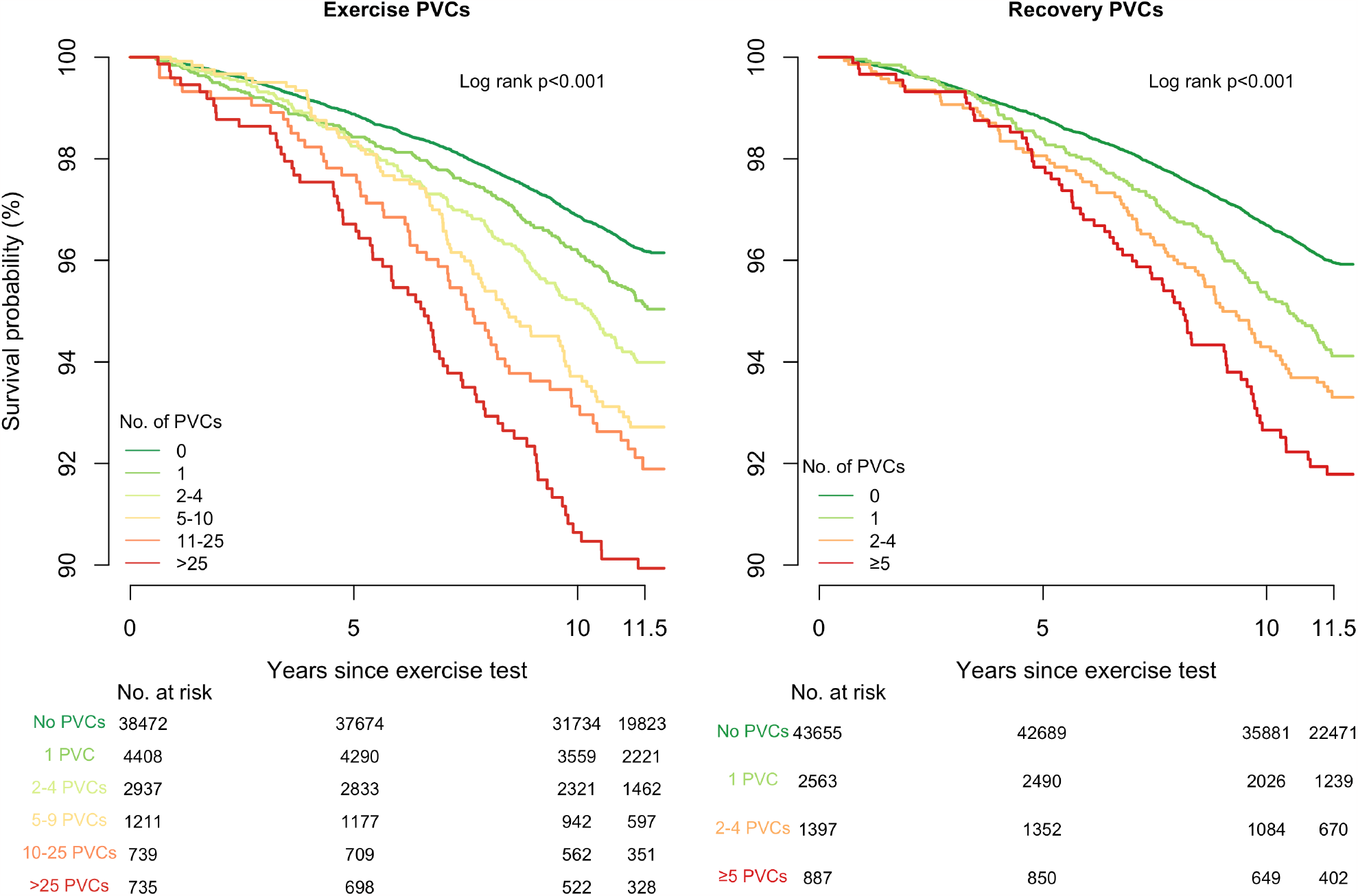
Survival Curves of Major Adverse Cardiovascular Events According to Different PVC Counts.

After adjusting for age, sex, and the number of heartbeats during exercise, all PVC counts were significantly associated with MACE, except for the lowest PVC count (1 PVC, Figure 3). The risk increased with PVC count with HRs ranging from: 1.2 (95% CI: 1.0 – 1.4, p = 0.022) for 2-4 PVCs to 1.8 (95% CI: 1.4 – 2.3, p < 0.001) for >25 PVCs. Regarding all-cause mortality, only the 2 highest PVC counts (11-25 and >25 PVCs) were associated with the endpoint (Supplemental Figure 3). For >25 PVCs, the HR was 1.5 (95% CI: 1.2 – 1.9, p = 0.002). After further adjusting using clinical and ECG variables, PVC count remained significantly associated with MACE and all-cause mortality with similar HRs (Figure 3, Supplemental Figure 3). In the same model, myocardial ischemia (ST depression) was not significantly associated with MACE or all-cause mortality (HR: 1.3, (95% CI: 0.8-1.9, p = 0.249), and HR: 1.4 (95% CI: 1.0-2.1, p = 0.055), respectively).

**Figure 3:**
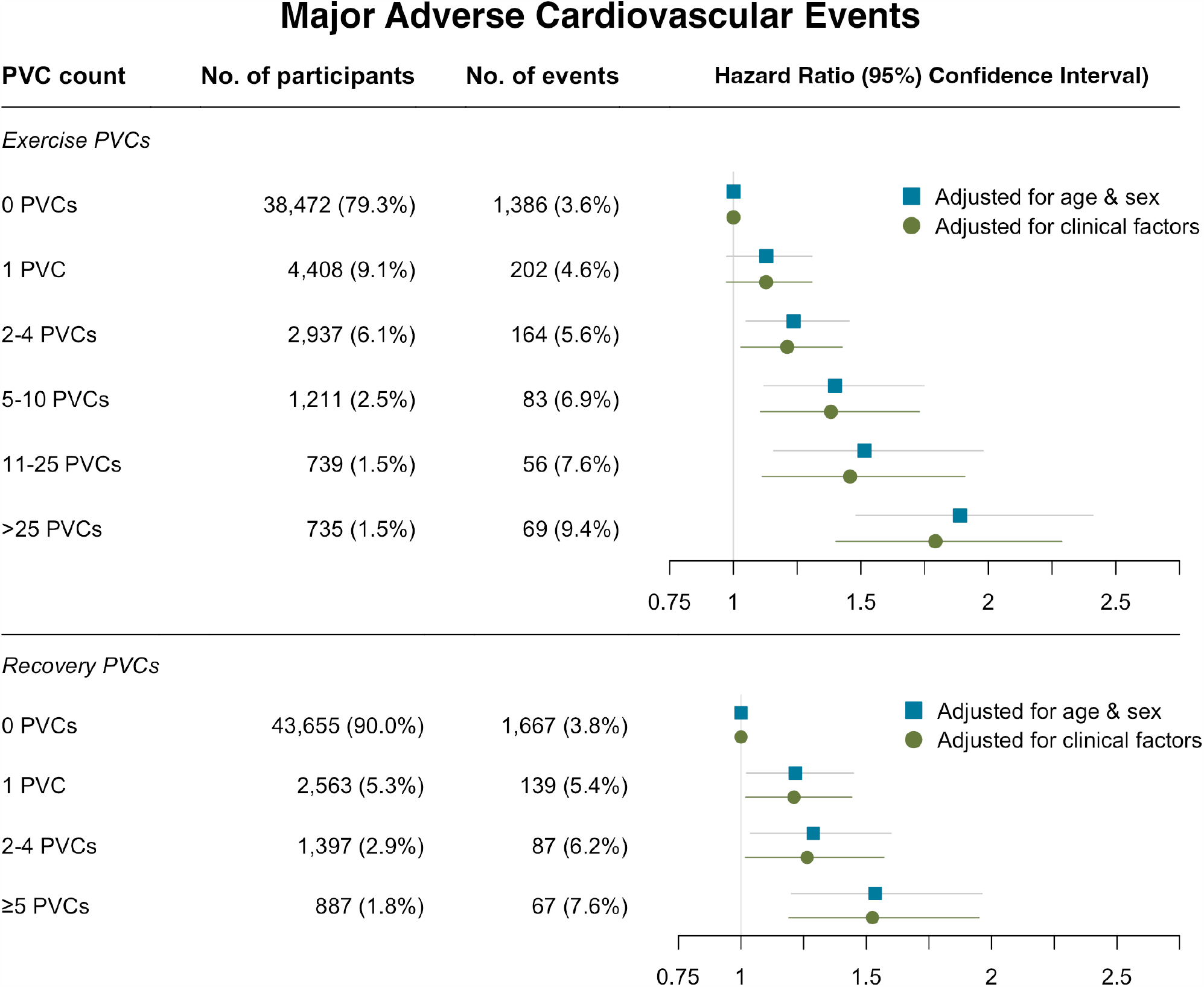
Adjusted Hazard Ratios of Major Adverse Cardiovascular Events According to PVC count. Hazard ratios (HRs) were adjusted for three models: (i) age sex, and no of beats (either during exercise or recovery); and (ii) clinical variables: age, sex, diabetes, hypertension, beta-blocker medication, smoking, LDL and HDL cholesterol, triglycerides, body mass index, QRS duration, QTc interval, ST depression (>0.1 mV), and no. of heartbeats during exercise and heart rate exercise (for exercise PVCs), or no. of heartbeats during recovery and heart rate recovery (for recovery PVCs). The HRs were statistically significant (p<0.05) for all PVC counts, except 1 PVC during exercise, and increased with increasing PVC count.

### PVCs during recovery from exercise

The MACE incidence was significantly higher in the group with PVCs: 3.8% (0 PVCs) vs. 5.4% (1 PVC, p < 0.001) and increased with PVC count among participants with PVCs (5.4% (1 PVC) to 7.6% (≥5 PVCs), p = 0.026). A similar trend was observed for all-cause mortality: the incidence increased from 4.0% (0 PVCs) to 5.5% (1 PVC, p < 0.001), and among participants with PVCs from 5.5% (1 PVC) to 8.5% (≥5 PVCs, p = 0.003). The Kaplan-Meier curves showed increasing event rates for both MACE and all-cause mortality with increasing PVC counts (p < 0.001, Figure 2, Supplemental Figure 2)).

After adjusting for age, sex, and the number of heartbeats during recovery, PVC count was significantly associated with MACE (adjusted HR ranging from: HR: 1.2 (95% CI: 1.0 – 1.4, p = 0.031) for 1 PVC to HR: 1.5 (95% CI: 1.2 – 1.9, p < 0.001) for ≥5 PVCs (Figure 3). Effect sizes were similar for the fully adjusted model (Figure 3). Only the highest PVC-count (≥5 PVCs) was associated with all-cause mortality: HR: 1.4 (95% CI: 1.1-1.8, p = 0.002, Supplemental Figure 3). After further adjusting using clinical and ECG variables, PVC count remained significantly associated with MACE and all-cause mortality with similar HRs (Figure 3, Supplemental Figure 3).

### Combining PVCs during exercise and recovery

A majority of the participants with repetitive PVCs during exercise also had PVCs during recovery: 457 (64%) from all participants with >25 PVCs during exercise had also ≥5 PVCs during recovery. This group comprised 52% of the subjects with ≥5 PVCs during recovery. After adjusting for clinical risk factors, the combination of >25 PVCs during exercise *and* ≥5 PVCs during recovery was associated with both MACE (HR: 1.8, 95% CI: 1.3 – 2.4, p < 0.001) and all-cause mortality (HR: 1.7, 95% CI: 1.2 – 2.2, p < 0.001).

### Association with additional study endpoints

The number of myocardial infarction, LTVA, and HF events were 1297 (2.7%), 276 (0.6%), and 691 (1.4%), respectively. There were 385 (0.8%) cardiovascular deaths. PVCs during exercise and recovery were both strongly associated with incident LTVAs, HF and cardiovascular mortality, but not with myocardial infarction (Supplemental Tables 5 and 6). The highest risk was observed for the highest PVC counts: For exercise, the fully adjusted HRs of >25 PVCs were: 3.4 (95% CI: 2.0 – 5.6, p < 0.001), 2.6 (95% CI: 1.9 – 3.7, p < 0.001), and 2.4 (95% CI: 1.5-3.8, p < 0.001), for LTVA, HF, and cardiovascular mortality, respectively. Similarly, the HRs of ≥5 recovery PVCs were: 2.2 (95% CI: 1.3-3.8, p = 0.003), 2.3 (95% CI: 1.6 – 3.1, p < 0.001), and 1.9 (95% CI: 1.1 – 3.1, p = 0.014) for LTVA, HF, and cardiovascular mortality, respectively.

### Prognostic value of high-grade & complex PVC rhythms

To compare our data with the recently published work from Refaat et al ^8^, we identified high-grade PVCs in 1655 (3.4%) and 748 (1.5%) participants, during exercise and recovery respectively. Both high grade PVCs and complex ventricular rhythms were associated with MACE independent of clinical risk and ECG factors (HR 1.7 (1.5-2.1), p<0.001; and HR 1.6 (1.2-2.1), p<0.001, for exercise and recovery, respectively, Figure 4). Only high-grade PVCs during exercise were associated with all-cause mortality (HR 1.2, 95% CI: 1.0-1.5, p = 0.024), but both markers were associated with cardiovascular mortality (HR 1.9, 95% CI 1.3-2.7, p<0.001, and HR 1.9, 95% CI 1.2-3.2, p = 0.016, Supplemental Table 7). In addition, we also observed that PVC couplets, triplets, and bigeminy were all associated with increased risk for MACE compared to PVC count alone (Figure 4).

**Figure 4:**
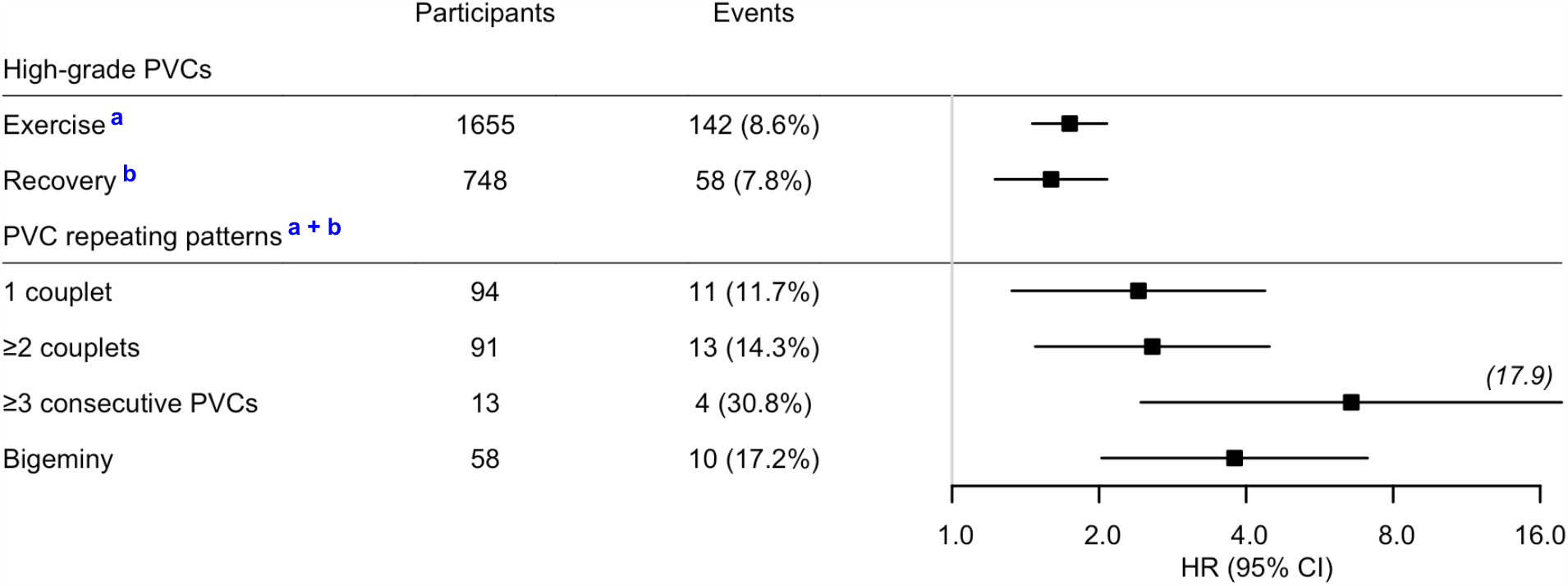
Adjusted Hazard Ratios of Major Adverse Cardiovascular Events According to Presence of Different PVC Patterns. Hazard ratios were adjusted for clinical and exercise test variables and were all statistically significant (P < 0.05). Clinical variables: age, sex, diabetes, hypertension, beta-blocker medication, LDL and HDL cholesterol, triglycerides, body mass index, QRS duration, QTc interval, ST depression (>0.1 mV), and exercise ECG variables: ^**a**^Total number of beats during exercise, and heart rate exercise; ^**b**^Total number of beats during recovery, and heart rate recovery. HRs were referenced against participants without PVCs during exercise (*a*), recovery (*b*), or both (*a* + *b*). PVC: premature ventricular complex. HR hazard ratio, CI confidence interval.

### Sensitivity analysis –high PVC burden at rest

High PVC burden at rest was observed in 1437 (3.0%) participants. The associations between exercise-induced PVCs and MACE and all-cause mortality remained significant with similar effect sizes as the main analysis (Supplemental Table 8 and 9).

## Discussion

To our knowledge, this is the largest population-based cohort study to date to investigate the prognostic value of exercise-induced PVCs in asymptomatic individuals without known cardiovascular disease. The main important findings are: (1) A very low PVC count (≥2 PVC during exercise or ≥1 PVC during recovery) was already significantly associated with MACE, independent of clinical risk factors, (2) PVC count during exercise and recovery are *both* associated with MACE, mortality, HF, LTVAs, and cardiovascular death, independent of clinical and exercise stress test factors; (3) there is a dose response relationship between PVC count and risk; and (4) complex PVCs rhythms are associated with higher risk compared to PVC count alone. This has important implications in the assessment of asymptomatic individuals particularly with the advent of wearable ECG technologies worn by otherwise healthy individuals during daily activity and exercise prompting cardiological review.

Recently, Refaat et al ^8^ investigated the prognostic value of PVC during exercise vs. recovery in 5,486 asymptomatic patients not suspected of having heart disease from the Lipid Research Clinics study. Compared to previous studies, they used a more inclusive definition of “high-grade PVCs” (>10 PVCs over 60-second interval, >2 consecutive PVCs including ventricular tachycardia, R-on-T PVCs, or multiform PVCs). Their data showed that high-grade PVCs during recovery were significantly associated with higher risk of cardiovascular mortality (adjusted HR: 1.7 (1.1-2.6); p = 0.02), but no associations were found for high-grade PVCs during exercise. Furthermore, high-grade PVCs (neither during exercise nor recovery) were not associated with all-cause mortality after adjusting for clinical variables. Our data from >48,000 participants of UK Biobank only partially support these findings. We found that high-grade PVCs during *exercise and recovery* were *both* associated with cardiovascular mortality, MACE, and all-cause mortality, using the same definition for high-grade PVCs and included similar clinical and exercise parameters as covariates. It is possible that conflicting differences in baseline characteristics between both populations e.g higher proportion of hyperlipidaemia patients and smokers compared to our cohort could explain these differences. Sensitivity analysis from Refaat et al ^8^ showed no link between PVC risk and hyperlipidemia, but other work investigating PVC load and aortic stiffness did find an association between the number of PVCs in 24h ambulatory monitoring ^22^. Another important difference between our and Refaat’s work is the exercise protocol. The activity performed by UK Biobank participants in this study was limited to either 35% or 50% of the maximum predicted workload, while subjects in Refaat‘s work ^8^ exercised at maximum workload. Plasma catecholamine levels have been linked with exercise intensity ^23,24^ and may mediate PVCs. It is possible that subjects in Refaat’s work have experienced higher catecholaminergic levels by exercising at maximum workload and that their exercise PVCs are more likely to reflect a physiological response to exercise compared to UK Biobank. However, this explanation may not be fully compatible with the “dose-response” observed in this work, which suggests that the risk for MACE and mortality increases with PVC count.

To our knowledge, this is the first study to document a “dose-response” association for exercise-induced PVCs in asymptomatic individuals without cardiovascular disease. Uniquely in this study we did not use one cut-off threshold for PVC burden but evaluated the risk for a wide range of PVC counts, during exercise and recovery. Little is known about the risk associated with low and intermediate PVC counts. Two previous studies have investigated the prognostic value of infrequent PVCs: Jouven et al ^9^ observed infrequent PVCs (<10% of all ventricular depolarizations, no consecutive PVCs) in 8.5% and 7.3% of a middle-aged male population during exercise and recovery, respectively. They found that cardiovascular and total mortality rates were higher for infrequent PVCs with respect to subjects without PVCs, however they only performed formal association studies for frequent PVCs (≥10% of all ventricular depolarizations). In the second study, Morshedi-Meibodi et al ^13^ investigated infrequent and frequent PVCs during exercise (< and ≥0.22/min of exercise) and found that both type of PVCs were significantly associated with mortality in 2885 participants from the Framingham Offspring Study during 15 years of follow-up (adjusted HR: 1.9 (1.2 – 2.8), and 1.7, (1.2 - 2.5), for infrequent and frequent PVCs, respectively). In our cohort, this cut-off value translates to between 1 and 2 PVCs during exercise. Both 1 and 2-4 PVC counts were not associated with all-cause mortality in this work.

### Pathophysiological Rationale

Previous studies in both patients and asymptomatic participants have suggested that PVCs during recovery carry a worse prognosis for (cardiovascular) mortality compared to PVCs during exercise^6,8^. This has fuelled the hypothesis that the prognostic value of exercise-induced PVCs might be linked to attenuated vagal reactivation during recovery, perhaps by reduced suppression of PVCs. Our data does support this hypothesis but indicates at least equal prognostic value for PVCs *during* exercise. PVCs during exercise may reflect increased norepinephrine levels during exercise in high-risk individuals, for example because they had to produce more effort during exercise compared to their healthier counterparts who may have more cardio-pulmonary reserve and skeletal muscle efficiency. The mechanisms of how PVCs during exercise and recovery are related to cardiovascular events remains to be further explored. The strong associations between PVC count and LTVA and HF implicates the existence of an arrhythmic substrate (i.e. fibrosis or ischemia), which enables re-entry and reduces cardiac function precipitating these events. The development of PVCs during exercise and recovery indicates the presence of this underlying substrate with PVCs arising due to the effects of subclinical ischaemia (reflected by ST depression on exercise) and adrenergic stress causing triggered ectopic activity which could be due to early afterdeporisations typically occurring in conditions of catecholamines, tissue injury, altered electrolytes, hypoxia, or acidosis^25^.

Increased PVCs during exercise may also reflect the presence of subclinical pathologic myocardial substrate or ischaemia. Although ST depression, was not associated with MACE nor mortality, it was increased for LVTA and HF. Ischaemia can promote optimal conditions for VAs & impair LV function through multiple mechanisms and it is well recognised to increase myocardial refractoriness to promote wavebreak and polymorphic VT^26,27^. However, the fact that single lead ST measurements with submaximal exercise could result in underestimation of ischaemia burden is a potential limitation in this study. Previously, using 2 bipolar leads (V5 and V5R) and *a maximal exercise* test frequent exercise-induced PVCs and ischemia were *independent predictors* of cardiovascular mortality with similar effect sizes in asymptomatic men^9^. In that dataset, ischemia was only present in 6% of PVC positive patients and 2% of our series, indicating other pathophysiological factors are operative. Combined with the lack of evidence to support an association between PVC count and myocardial infarction, findings seem to suggest that exercise and recovery PVCs are not necessarily related to myocardial ischemia^9^.

The relationship between PVCs recorded during rest and exercise remains to be further investigated. Previously, we have shown that high PVC burden at rest was associated with several cardiovascular outcomes^28^. Results from the sensitivity analyses in the current work seems to suggest that the associations between exercise-induced PVCs and MACE and all-cause mortality were independent from PVC burden at rest before exercise. However, the resting ECGs (recorded pre-exercise) were of very short duration (15s) and therefore likely to have provided very limited sensitivity to detect PVCs as their occurrence is highly variable.

### Limitations

Our study benefited from several important advantages including access to the largest and most detailed dataset of PVC data from asymptomatic data without known cardiovascular disease from a population-based cohort, combined with long-term follow-up. Nevertheless, several limitations, apart from the limited sensitivity to detect myocardial ischemia need to be acknowledged. First, there is a “healthy volunteer” selection bias in the UK Biobank with the participants being older and healthier than the general population. Third, the study cohort is predominantly (>90%) of white British ancestry, which may limit the generalisability of our findings in under-represented ethnicities. However, the study is not likely to be flawed by an important co-morbidity burden. Finally, no contemporaneous cardiac imaging data were available in this study (imaging data in UK Biobank were recorded 5-10 years after the exercise stress test). This requires future evaluation to investigate the underlying mechanisms.

### Clinical Perspectives

In the recent years there has been a widespread use of wearable cardiac rhythm monitors, many of which are used during exercise. This has enabled detection of PVCs in asymptomatic individuals at larger scale and is expected to result in more consultation requests from concerned patients whose monitors detect PVCs. PVC count during exercise and recovery predicts MACE and all-cause mortality independent of standard CV risk factors. Consequently, clinicians need to be aware of the measurable prognostic significance of these PVCs to optimally assess and manage CV risk in these otherwise asymptomatic individuals to prevent life threatening events.

### Conclusions

There is a “dose response” of PVC count during exercise and recovery for the association with MACE and all-cause mortality independent of standard CV risk factors. In addition, complex PVCs have important prognostic significance. This has major implications for risk stratification, population screening and interpretation of wearable ECG rhythm readouts in the general population provoking increased referrals for cardiology review.

## Data Availability

All data produced in the present study will be made available upon reasonable request to UK Biobank

## Funding

This study was supported by the MRC grant MR/N025083/1. PDL is supported by UCL/UCLH Biomedicine NIHR. AT and PBM and AT acknowledge the NIHR Cardiovascular Biomedical Research Centre at Barts and QMUL, UK. JR acknowledges the “María Zambrano” fellowship support from the European Union-NextGenerationEU. AD and SVD also acknowledge support from the Wellcome Trust (223100/Z/21/Z).

## Conflict of interest

None.

## Data availability statement

Anonymised data and materials generated in this work will be returned to UK Biobank and can be accessed upon request.

## Notes

### Competing Interest Statement

The authors have declared no competing interest.

### Author Declarations

The study has extensive baseline and follow-up clinical, biochemical, and outcome measures and ap-proval from the North West Multi-Centre Research Ethics Committee. All participants provided in-formed consent at the time of enrolment for their data to be linked to the health record systems.

