## Supplemental Figures for "Prognostic Significance of Different Ventricular Ectopic Burdens During Exercise in Asymptomatic UK Biobank Subjects"

### Figure S1  Distributions of PVC Counts

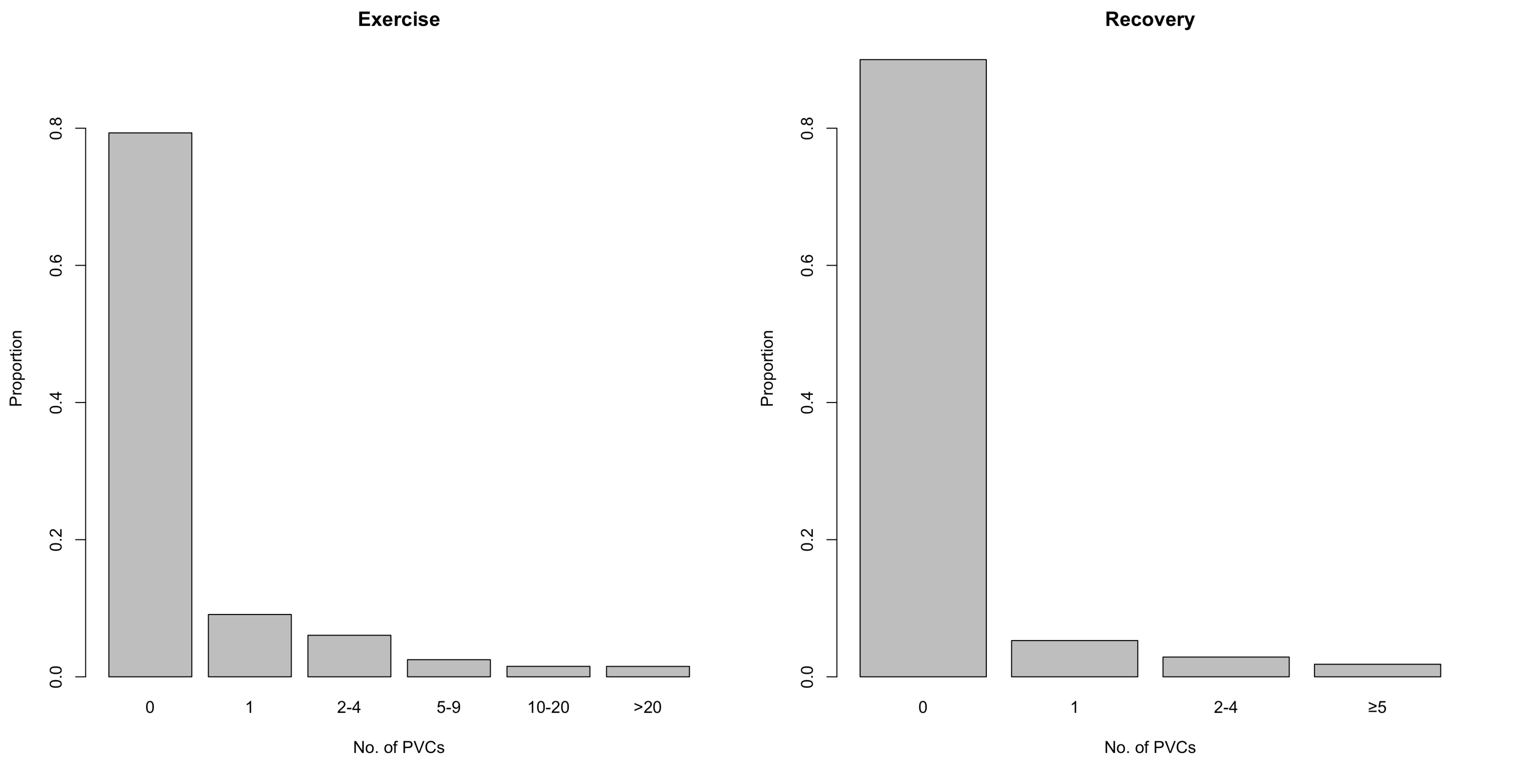

### Figure S2 Survival Curves of All-Cause Mortality According to Different PVC Counts

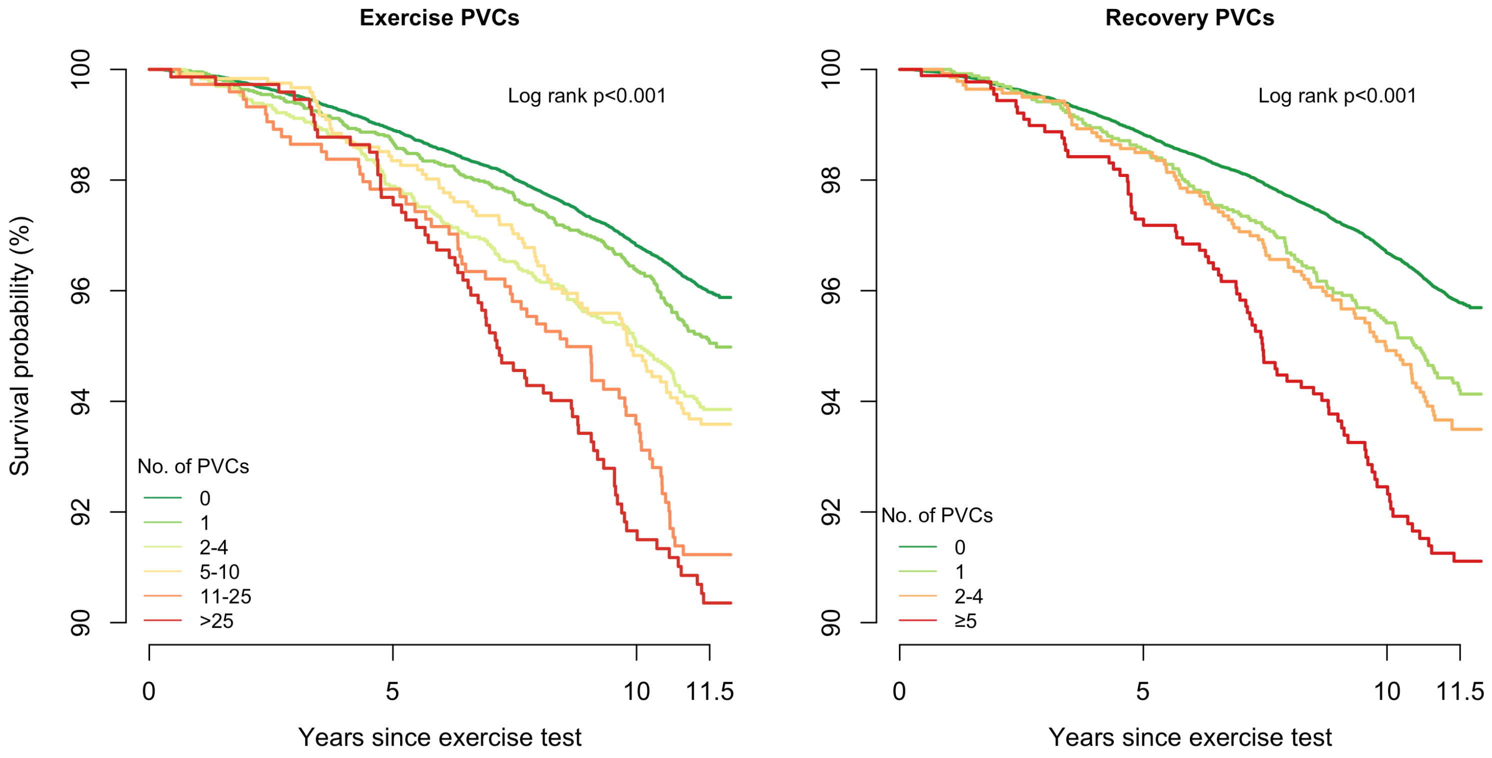


No. at risk (Exercise PVCs)

| | 0 | 5 | 10 | 11.5 |
|---|---|---|---|---|
| No PVCs | 38472 | 38051 | 32597 | 20460 |
| 1 PVC | 4408 | 4350 | 3676 | 2318 |
| 2-4 PVCs | 2937 | 2875 | 2418 | 1547 |
| 5-9 PVCs | 1211 | 1191 | 993 | 633 |
| 10-25 PVCs | 739 | 723 | 595 | 380 |
| >25 PVCs | 735 | 718 | 568 | 357 |

No. at risk (Recovery PVCs)

| | 0 | 5 | 10 | 11.5 |
|---|---|---|---|---|
| No PVCs | 43655 | 43143 | 36910 | 23247 |
| 1 PVC | 2563 | 2526 | 2109 | 1309 |
| 2-4 PVCs | 1397 | 1376 | 1135 | 708 |
| ≥5 PVCs | 887 | 863 | 693 | 431 |

**Figure S3 Adjusted Hazard Ratios of All-Cause Mortality According to PVC count**

#### All-Cause Mortality

| PVC count | No. of participants | No. of events | Hazard Ratio (95%) Confidence Interval) |
|---|---|---|---|
| *Exercise PVCs* | | | |
| 0 PVCs | 38,472 (79.3%) | 1,386 (3.6%) | |
| 1 PVC | 4,408 (9.1%) | 202 (4.6%) | |
| 2-4 PVCs | 2,937 (6.1%) | 164 (5.6%) | |
| 5-10 PVCs | 1,211 (2.5%) | 83 (6.9%) | |
| 11-25 PVCs | 739 (1.5%) | 56 (7.6%) | |
| >25 PVCs | 735 (1.5%) | 69 (9.4%) | |

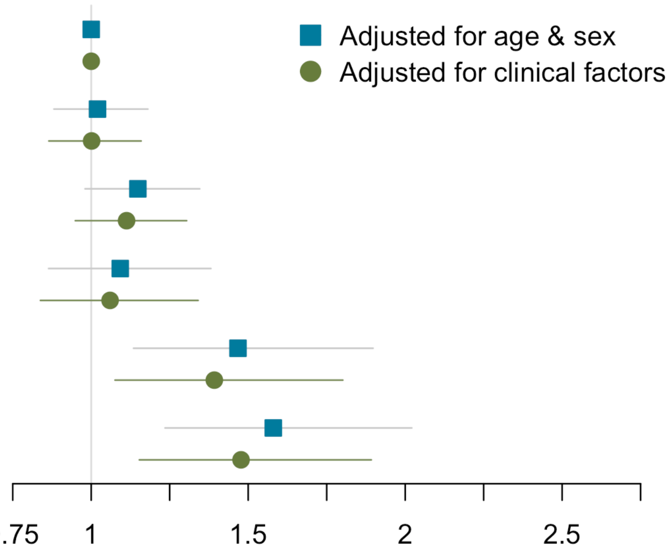


| PVC count | No. of participants | No. of events | Hazard Ratio (95%) Confidence Interval) |
|---|---|---|---|
| *Recovery PVCs* | | | |
| 0 PVCs | 43,655 (90.0%) | 1,667 (3.8%) | |
| 1 PVC | 2,563 (5.3%) | 139 (5.4%) | |
| 2-4 PVCs | 1,397 (2.9%) | 87 (6.2%) | |
| ≥5 PVCs | 887 (1.8%) | 67 (7.6%) | |

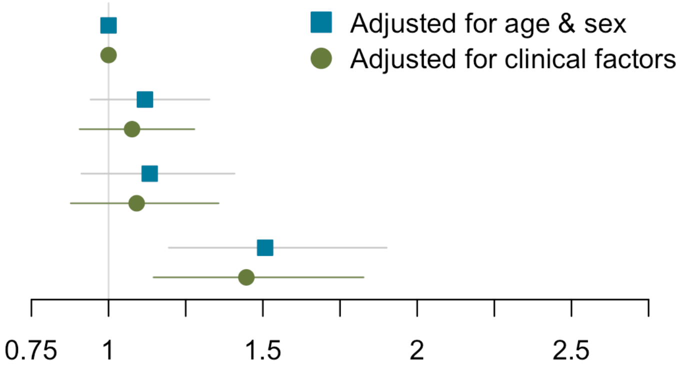


Hazard ratios (HRs) were adjusted for three models: (i) age sex, and no of beats (either during exercise or recovery); and (ii) clinical variables: age, sex, diabetes, hypertension, beta-blocker medication, smoking, LDL and HDL cholesterol, triglycerides, body mass index, QRS duration, QTc interval, ST depression (>0.1 mV), and no. of heartbeats during exercise and heart rate exercise (for exercise PVCs), or no. of heartbeats during recovery and heart rate recovery (for recovery PVCs). For exercise PVCs, HRs were only statistically significant ($p<0.05$) for counts with >10 PVCs. For recovery PVCs, HRs were only statistically significant for ≥5 PVCs.
