## Supplemental Tables for "Prognostic Significance of Different Ventricular Ectopic Burdens During Exercise in Asymptomatic UK Biobank Subjects"

**Supplemental Table 1:** Performance of the machine learning method for PVC detection

| **Training data (N=723 ECGs)** | | | | | | |
|---|---|---|---|---|---|---|
| Overall Accuracy (%) | **96.7%** | | | | | |
| PVC count category | 0 | 1 | 2 – 4 | 5 – 10 | 11 – 25 | >25 |
| N (%) | 154 (21.3%) | 41 (5.7%) | 55 (7.6%) | 47 (6.5%) | 67 (9.2%) | 359 (49.7%) |
| Sensitivity (%) | 97.4 | 92.7 | 94.5 | 93.6 | 98.5 | 97.2 |
| Positive Predictive Value (%) | 99.3 | 88.4 | 91.2 | 93.6 | 86.8 | 100.0 |

| **Validation data (N=1500 ECGs)** | | | | | | |
|---|---|---|---|---|---|---|
| Overall Accuracy (%) | **98.5%** | | | | | |
| PVC count category | 0 | 1 | 2 – 4 | 5 – 10 | 11 – 25 | >25 |
| N (%) | 1119 (74.6%) | 169 (11.2%) | 101 (6.7%) | 50 (3.3%) | 35 (2.3%) | 26 (1.7%) |
| Sensitivity (%) | 99.0 | 98.2 | 97.0 | 98.0 | 91.4 | 96.0 |
| Positive Predictive Value (%) | 99.8 | 94.3 | 96.0 | 96.0 | 94.1 | 92.6 |

*1 ECG per participant, PVC = premature ventricular contraction*

**Supplementary Table 2:** Codes for diagnoses of cardiovascular events used for excluding participant with prevalent cardiovascular disease at baseline

| Subgroup | ICD9 codes | ICD10 codes | Definition |
|---|---|---|---|
| Ischemic Heart Disease | 411.1 | I20 | Unstable angina |
| | | I21 | Acute myocardial infarction |
| | 410.11 | I21.0 | Acute transmural myocardial infarction of anterior wall |
| | 410.41 | I21.1 | Acute transmural myocardial infarction of inferior wall |
| | 410.81 | I21.2 | Acute transmural myocardial infarction of other sites |
| | 410.91 | I21.3 | Acute transmural myocardial infarction of unspecified site |
| | 410.71 | I21.4 | Acute subendocardial myocardial infarction |
| | 410.91 | I21.9 | Acute myocardial infarction, unspecified |
| | | I22 | Subsequent myocardial infarction |
| | 410.01/410.11 | I22.0 | Subsequent myocardial infarction of anterior wall |
| | 410.21/410.31/410.41 | I22.1 | Subsequent myocardial infarction of inferior wall |
| | 410.51/410.61/410.81 | I22.8 | Subsequent myocardial infarction of other sites |
| | 410.91 | I22.9 | Subsequent myocardial infarction of unspecified site |
| | | I24 | Other acute ischaemic heart diseases |
| | | I24.0 | Coronary thrombosis not resulting in myocardial infarction |
| | 411.89 | I24.8 | Other forms of acute ischaemic heart disease |
| | 411.89 | I24.9 | Acute ischaemic heart disease, unspecified |
| | | I25 | Chronic ischaemic heart disease |
| | 429.2 | I25.0 | Atherosclerotic cardiovascular disease, so described |
| | 414.0 | I25.1 | Atherosclerotic heart disease |
| | | I25.2 | Old myocardial infarction |
| | 414.10/414.19 | I25.3 | Aneurysm of heart |
| | | I25.4 | Coronary artery aneurysm |
| | 414.8 | I25.5 | Ischaemic cardiomyopathy |
| | 414.8 | I25.6 | Silent myocardial ischaemia |
| | | I25.8 | Other forms of chronic ischaemic heart disease |
| | 414.8/414.9 | I25.9 | Chronic ischaemic heart disease, unspecified |
| Cardiomyopathies | | I42 | Cardiomyopathy |
| | 425.4 | I42.0 | Dilated cardiomyopathy |
| | 425.11 | I42.1 | Obstructive hypertrophic cardiomyopathy |
| | 425.18 | I42.2 | Other hypertrophic cardiomyopathy |
| | 425 | I42.3 | Endomyocardial (eosinophilic) disease |
| | 425.3 | I42.4 | Endocardial fibroelastosis |
| | 425.4 | I42.5 | Other restrictive cardiomyopathy |
| | 425.5 | I42.6 | Alcoholic cardiomyopathy |
| | 425.9 | I42.7 | Cardiomyopathy due to drugs and other external agents |
| | 425.2/425.4 | I42.8 | Other cardiomyopathies |
| | 425.4/425.9 | I42.9 | Cardiomyopathy, unspecified |
| | 425.8 | I43 | Cardiomyopathy in diseases classified elsewhere |
| | | I43.0 | Cardiomyopathy in infectious and parasitic diseases classified elsewhere |
| | 425.7 | I43.1 | Cardiomyopathy in metabolic diseases |
| | 425.7 | I43.2 | Cardiomyopathy in nutritional diseases |
| | 425.8 | I43.8 | Cardiomyopathy in other diseases classified elsewhere |
| Heart Failure | | I50 | Heart failure |
| | 428.0 | I50.0 | Congestive heart failure |
| | 428.1 | I50.1 | Left ventricular failure |
| | 428.0/428.9 | I50.9 | Heart failure, unspecified |
| Atrial Arrhythmia | 427.3 | I48 | Atrial fibrillation |
| | 427.31 | I48.0 | Paroxysmal atrial fibrillation |
| | 427.31 | I48.1 | Persistent atrial fibrillation |
| | 427.31 | I48.2 | Chronic atrial fibrillation |
| | 427.32 | I48.3 | Typical atrial flutter |
| | 427.32 | I48.4 | Atypical atrial flutter |
| | | I48.9 | Atrial fibrillation and atrial flutter, unspecified |
| Ventricular Arrhythmia | 427.1 | I47.2 | Ventricular tachycardia |
| | | I49.0 | Ventricular fibrillation and flutter |
| Significant conduction disease | | I44.1 | Atrioventricular block, second degree |
| | | I44.2 | Atrioventricular block, complete |
| Arrhythmia general | 427.9 | I49.9 | Cardiac arrhythmia, unspecified |
| Cardiac Arrest | 427.5 | I46.1 | Sudden cardiac death, so described |
| | 427.5 | I46.9 | Cardiac arrest, unspecified |
| SCD | 427.5 | I46.1 | Sudden cardiac death, so described |
| TIA | | G45 | Transient cerebral ischaemic attacks and related syndromes |
| | | G45.0 | Vertebro-basilar artery syndrome |
| | | G45.3 | Amaurosis fugax |
| | | G45.8 | Other transient cerebral ischaemic attacks and related |
| | | G45.9 | Transient cerebral ischaemic attack, unspecified |
| Ischaemic Stroke | | I63 | Cerebral infarction |
| | 434.91 | I63.0 | Cerebral infarction due to thrombosis of precerebral arteries |
| | 434.91 | I63.1 | Cerebral infarction due to embolism of precerebral arteries |
| | 434.91 | I63.2 | Cerebral infarction due to unspecified occlusion or stenosis of precerebral arteries |
| | 434.01 | I63.3 | Cerebral infarction due to thrombosis of cerebral arteries |
| | 434.11 | I63.4 | Cerebral infarction due to embolism of cerebral arteries |
| | 434.91 | I63.5 | Cerebral infarction due to unspecified occlusion or stenosis of cerebral arteries |
| | | I63.8 | Other cerebral infarction |
| | 434.91 | I63.9 | Cerebral infarction, unspecified |
| Haemorrhagic stroke | | I61 | Intracerebral haemorrhage |
| | | I61.0 | Intracerebral haemorrhage in hemisphere, subcortical |
| | | I61.1 | Intracerebral haemorrhage in hemisphere, cortical |
| | | I61.2 | Intracerebral haemorrhage in hemisphere, unspecified |
| | | I61.3 | Intracerebral haemorrhage in brain stem |
| | | I61.4 | Intracerebral haemorrhage in cerebellum |
| | | I61.5 | Intracerebral haemorrhage, intraventricular |
| | | I61.6 | Intracerebral haemorrhage, multiple localised |
| | | I61.8 | Other intracerebral haemorrhage |
| | | I61.9 | Intracerebral haemorrhage, unspecified |
| Unspecified stroke | 436 | I64 | Stroke, not specified as haemorrhage or infarction |
| rtic/Peripheral vascular disease | | I70 | Atherosclerosis |
| | | I70.0 | Atherosclerosis of aorta |
| | | I70.00 | Atherosclerosis of aorta (without gangrene) |
| | | I70.01 | Atherosclerosis of aorta (with gangrene) |
| | | I70.2 | Atherosclerosis of arteries of the extremities |
| | | I70.20 | Atherosclerosis of arteries of extremities (without gangrene) |
| | | I70.21 | Atherosclerosis of arteries of extremities (with gangrene) |
| | | I70.8 | Atherosclerosis of other arteries |
| | | I70.80 | Atherosclerosis of other arteries (without gangrene) |
| | | I73 | Other peripheral vascular diseases |
| | | I73.0 | Raynaud's syndrome |
| | | I73.1 | Thromboangiitis obliterans [Buerger] |
| | | I73.8 | Other specified peripheral vascular diseases |
| | | I73.9 | Peripheral vascular disease, unspecified |
| | | I74 | Arterial embolism and thrombosis |

| | | |
|---|---|---|
| | I74.0 | Embolism and thrombosis of abdominal aorta |
| | I74.1 | Embolism and thrombosis of other and unspecified parts of aorta |
| | I74.2 | Embolism and thrombosis of arteries of the upper extremities |
| | I74.3 | Embolism and thrombosis of arteries of the lower extremities |
| | I74.4 | Embolism and thrombosis of arteries of extremities, unspecified |
| | I74.5 | Embolism and thrombosis of iliac artery |
| | I74.8 | Embolism and thrombosis of other arteries |
| | I74.9 | Embolism and thrombosis of unspecified artery |
| Other vascular disease | I71 | Aortic aneurysm and dissection |
| | I71.0 | Dissection of aorta [any part] |
| | I71.1 | Thoracic aortic aneurysm, ruptured |
| | I71.2 | Thoracic aortic aneurysm, without mention of rupture |
| | I71.3 | Abdominal aortic aneurysm, ruptured |
| | I71.4 | Abdominal aortic aneurysm, without mention of rupture |
| | I71.5 | Thoracoabdominal aortic aneurysm, ruptured |
| | I71.6 | Thoracoabdominal aortic aneurysm, without mention of rupture |
| | I71.8 | Aortic aneurysm of unspecified site, ruptured |
| | I71.9 | Aortic aneurysm of unspecified site, without mention of rupture |
| Hypertensive heart disease | I11 | Hypertensive heart disease |
| | I11.0 | Hypertensive heart disease with (congestive) heart failure |
| | I11.9 | Hypertensive heart disease without (congestive) heart failure |
| -rheumatic valvular heart disease | I34 | Nonrheumatic mitral valve disorders |
| | I34.0 | Mitral (valve) insufficiency |
| | I34.1 | Mitral (valve) prolapse |
| | I34.2 | Nonrheumatic mitral (valve) stenosis |
| | I34.8 | Other nonrheumatic mitral valve disorders |
| | I34.9 | Nonrheumatic mitral valve disorder, unspecified |
| | I35 | Nonrheumatic aortic valve disorders |
| | I35.0 | Aortic (valve) stenosis |
| | I35.1 | Aortic (valve) insufficiency |
| | I35.2 | Aortic (valve) stenosis with insufficiency |
| | I35.8 | Other aortic valve disorders |
| | I35.9 | Aortic valve disorder, unspecified |
| | I36 | Nonrheumatic tricuspid valve disorders |
| | I36.0 | Nonrheumatic tricuspid (valve) stenosis |
| | I36.1 | Nonrheumatic tricuspid (valve) insufficiency |
| | I36.8 | Other nonrheumatic tricuspid valve disorders |
| | I36.9 | Nonrheumatic tricuspid valve disorder, unspecified |
| | I37 | Pulmonary valve disorders |
| | I37.0 | Pulmonary valve stenosis |
| | I37.1 | Pulmonary valve insufficiency |
| | I37.2 | Pulmonary valve stenosis with insufficiency |
| | I37.8 | Other pulmonary valve disorders |
| | I37.9 | Pulmonary valve disorder, unspecified |
| Congenital heart disease | Q20 | Congenital malformations of cardiac chambers and connexions |
| | Q20.0 | Common arterial trunk |
| | Q20.1 | Double outlet right ventricle |
| | Q20.2 | Double outlet left ventricle |
| | Q20.3 | Discordant ventriculoarterial connexion |
| | Q20.4 | Double inlet ventricle |
| | Q20.5 | Discordant atrioventricular connexion |
| | Q20.6 | Isomerism of atrial appendages |
| | Q20.8 | Other congenital malformations of cardiac chambers and connexions |
| | Q20.9 | Congenital malformation of cardiac chambers and connexions, unspecified |
| | Q21 | Congenital malformations of cardiac septa |
| | Q21.0 | Ventricular septal defect |
| | Q21.1 | Atrial septal defect |
| | Q21.2 | Atrioventricular septal defect |
| | Q21.3 | Tetralogy of Fallot |
| | Q21.4 | Aortopulmonary septal defect |
| | Q21.8 | Other congenital malformations of cardiac septa |
| | Q21.9 | Congenital malformation of cardiac septum, unspecified |
| | Q22 | Congenital malformations of pulmonary and tricuspid valves |
| | Q22.1 | Congenital pulmonary valve stenosis |
| | Q22.2 | Congenital pulmonary valve insufficiency |
| | Q22.4 | Congenital tricuspid stenosis |
| | Q22.5 | Ebstein's anomaly |
| | Q22.8 | Other congenital malformations of tricuspid valve |
| | Q22.9 | Congenital malformation of tricuspid valve, unspecified |
| | Q23 | Congenital malformations of aortic and mitral valves |
| | Q23.0 | Congenital stenosis of aortic valve |
| | Q23.1 | Congenital insufficiency of aortic valve |
| | Q23.2 | Congenital mitral stenosis |
| | Q23.3 | Congenital mitral insufficiency |
| | Q23.4 | Hypoplastic left heart syndrome |
| | Q23.8 | Other congenital malformations of aortic and mitral valves |
| | Q23.9 | Congenital malformation of aortic and mitral valves, unspecified |
| | Q24 | Other congenital malformations of heart |
| | Q24.0 | Dextrocardia |
| | Q24.1 | Levocardia |
| | Q24.3 | Pulmonary infundibular stenosis |
| | Q24.4 | Congenital subaortic stenosis |
| | Q24.5 | Malformation of coronary vessels |
| | Q24.6 | Congenital heart block |
| | Q24.8 | Other specified congenital malformations of heart |
| | Q24.9 | Congenital malformation of the heart, unspecified |
| Myocarditis | B33.2 | Viral carditis |
| | I40.0 | Acute myocarditis |
| | I40.1 | Isolated myocarditis |
| | I40.8 | Other acute myocarditis |
| | I40.9 | Acute myocarditis, unspecified |
| | I41.1 | Myocarditis in viral diseases classified elsewhere |
| | I41.2 | Myocarditis in other infectious and parasitic diseases classified elsewhere |
| | I41.8 | Myocarditis in other diseases classified elsewhere |
| | I51.4 | Myocarditis, unspecified |

**Supplementary Table 3:** Codes for Primary and Secondary Outcomes

| **Myocardial Infarction** | |
|---|---|
| *ICD10 codes* | |
| I21 | Acute myocardial infarction |
| I21.0 | Acute transmural myocardial infarction of anterior wall |
| I21.1 | Acute transmural myocardial infarction of inferior wall |
| I21.2 | Acute transmural myocardial infarction of other sites |
| I21.3 | Acute transmural myocardial infarction of unspecified site |
| I21.4 | Acute subendocardial myocardial infarction |
| I21.9 | Acute myocardial infarction, unspecified |
| I22 | Subsequent myocardial infarction |
| I22.0 | Subsequent myocardial infarction of anterior wall |
| I22.1 | Subsequent myocardial infarction of inferior wall |
| I22.8 | Subsequent myocardial infarction of other sites |
| I22.9 | Subsequent myocardial infarction of unspecified site |
| I23 | Certain current complications following acute myocardial infarction |
| I23.0 | Haemopericardium as current complication following acute myocardial infarction |
| I23.1 | Atrial septal defect as current complication following acute myocardial infarction |
| I23.2 | Ventricular septal defect as current complication following acute myocardial infarction |
| I23.3 | Rupture of cardiac wall without haemopericardium as current complication following acute myocardial infarction |
| I23.4 | Rupture of chordae tendineae as current complication following acute myocardial infarction |
| I23.5 | Rupture of papillary muscle as current complication following acute myocardial infarction |
| I23.6 | Thrombosis of atrium , auricular appendage and ventricle as current complications following acute myocardial infarction |
| I23.8 | Other current complications following acute myocardial infarction |
| *Operative procedures* | |
| K40 | Saphenous vein graft replacement of coronary artery |
| K40.1 | Saphenous vein graft replacement of one coronary artery |
| K40.2 | Saphenous vein graft replacement of two coronary arteries |
| K40.3 | Saphenous vein graft replacement of three coronary arteries |
| K40.4 | Saphenous vein graft replacement of four or more coronary arteries |
| K40.9 | Unspecified saphenous vein graft replacement of coronary artery |
| K41 | Other autograft replacement of coronary artery |
| K41.1 | Autograft replacement of one coronary artery NEC |
| K41.2 | Autograft replacement of two coronary arteries NEC |
| K41.3 | Autograft replacement of three coronary arteries NEC |
| K41.4 | Autograft replacement of four or more coronary arteries NEC |
| K42 | Allograft replacement of coronary artery |
| K42.4 | Allograft replacement of four or more coronary arteries |
| K44 | Other replacement of coronary artery |
| K44.1 | Replacement of coronary arteries using multiple methods |
| K44.2 | Revision of replacement of coronary artery |
| K44.9 | Unspecified other replacement of coronary artery |
| K45 | Connection of thoracic artery to coronary artery |
| K45.1 | Double anastomosis of mammary arteries to coronary arteries |
| K45.2 | Double anastomosis of thoracic arteries to coronary arteries NEC |
| K45.3 | Anastomosis of mammary artery to left anterior descending coronary artery |
| K45.4 | Anastomosis of mammary artery to coronary artery NEC |
| K45.5 | Anastomosis of thoracic artery to coronary artery NEC |
| K45.6 | Revision of connection of thoracic artery to coronary artery |
| K45.8 | Other specified connection of thoracic artery to coronary artery |
| K45.9 | Unspecified connection of thoracic artery to coronary artery |
| K49 | Transluminal balloon angioplasty of coronary artery |
| K49.1 | Percutaneous transluminal balloon angioplasty of one coronary artery |
| K49.2 | Percutaneous transluminal balloon angioplasty of multiple coronary arteries |
| K49.3 | Percutaneous transluminal balloon angioplasty of bypass graft of coronary artery |
| K49.4 | Percutaneous transluminal cutting balloon angioplasty of coronary artery |
| K49.8 | Other specified transluminal balloon angioplasty of coronary artery |
| K49.9 | Unspecified transluminal balloon angioplasty of coronary artery |
| K50 | Other therapeutic transluminal operations on coronary artery |
| K50.1 | Percutaenous transluminal laser coronary angioplasty |
| K50.2 | Percutaneous transluminal coronary thrombolysis using streptokinase |
| K50.3 | Percutaneous transluminal injection of therapeutic substance into coronary artery NEC |
| K50.4 | Percutaenous transluminal atherectomy of coronary artery |
| K50.8 | Other specified other therapeutic transluminal operations on coronary artery |
| K50.9 | Unspecified other therapeutic transluminal operations on coronary artery |
| K75 | Percutaneous transluminal balloon angioplasty and insertion of stent into coronary artery |
| K75.1 | Percutaneous transluminal balloon angioplasty and insertion of 1-2 drug-eluting stents into coronary artery |
| K75.2 | Percutaneous transluminal balloon angioplasty and insertion of 3 or more drug-eluting stents into coronary artery |
| K75.3 | Percutaneous transluminal balloon angioplasty and insertion of 1-2 stents into coronary artery |
| K75.4 | Percutaneous transluminal balloon angioplasty and insertion of 3 or more stents into coronary artery NEC |
| K75.8 | Other specified percutaenous transluminal balloon angioplasty and insertion of stent into coronary artery |
| K75.9 | Unspecified percutaenous transluminal balloon angioplasty and insetion of stent into coronary artery |
| K59.6 | Implantation of cardioverter defibrillator using three electrode leads |
| K61.7 | Implantation of biventricular cardiac pacemaker system |
| K60.7 | Implantation of intravenous biventricular cardiac pacemaker system |

| **Heart Failure** | |
|---|---|
| *ICD10 codes* | |
| I13.0 | Hypertensive heart and renal disease with both (congestive) heart failure |
| I13.2 | Hypertensive heart and renal disease with both (congestive) heart failure and renal failure |
| I50 | Heart failure |
| I50.0 | Congestive heart failure |
| I50.1 | Left ventricular failure |
| I50.9 | Heart failure, unspecified |
| *ICD9 codes* | |
| 4280 | Congestive heart failure |
| 4281 | Left heart failure |
| 4289 | Heart failure, unspecified |
| *Operative procedures* | |
| K59.6 | Implantation of cardioverter defibrillator using three electrode leads |
| K61.7 | Implantation of biventricular cardiac pacemaker system |
| K60.7 | Implantation of intravenous biventricular cardiac pacemaker system |

| **Life threatening ventricular tachycardia** |
|---|

| | |
|---|---|
| *ICD10 codes* | |
| I47.2 | Ventricular tachycardia |
| I49.0 | Ventricular fibrillation and flutter |
| I46.0 | Cardiac arrest with successful resuscitation |
| I46.1 | Sudden cardiac death, so described |
| I46.9 | Cardiac arrest, unspecified |
| *Operative procedures* | |
| I47.0 | Re-entry ventricular arrhythmia |
| K57.6 | Percutaneous transluminal ablation of ventricular wall |
| K64.1 | Percutaneous radiofrequency ablation of epicardium |
| X50.3 | Advanced cardiac pulmonary resuscitation |
| X50.4 | External ventricular defibrillation |

**ICD Implant (included for major adverse cardiovascular event endpoint)**

| **OPCS4** | **Definition (41200)** |
|---|---|
| K59 | Cardioverter defibrillator introduced through vein |
| K59.1 | Implantation of cardioverter defibrillator using one electrode lead |
| K59.2 | Implantation of cardioverter defibrillator using two electrode leads |
| K59.3 | Resiting of lead of cardioverter defibrillator |
| K59.4 | Renewal of cardioverter defibrillator |
| K59.6 | Implantation of cardioverter defibrillator using three electrode leads |
| K59.8 | Other specified cardioverter defibrillator introduced through the vein |
| K59.9 | Unspecified cardioverter defibrillator introduced through the vein |
| K72 | Other cardioverter defibrillator |
| K72.1 | Implantation of subcutaenous cardioverter defibrillator |
| K72.3 | Renewal of subcutaneous cardioverter defibrillator |

**Supplemental Table 4:** Definition of hypertension and corresponding variables in UK Biobank

| Variable | UK Biobank Data Field |
|---|---|
| Systolic blood pressure > 140 mmHg | 4080, 93 |
| Diastolic blood pressure > 90 mmHg | 4079, 94 |
| Taking medication for high blood pressure | 6177, 6153 |
| High blood pressure diagnosed by doctor | 6150 |
| Age high blood pressure diagnosed | 2966 |

Supplemental Table 5: Adjusted Hazard Ratios of Myocardial Infarction, Heart Failure, and Life threatening Ventricular Arrhythmia to PVC count during exercise

| | | Myocardial Infarction | | Heart Failure | | Life threatening Ventricular Arrhythmia | | Cardiovascular Death | |
|---|---|---|---|---|---|---|---|---|---|
| | | HR (95% CI) | *P* Value | HR (95% CI) | *P* Value | HR (95% CI) | *P* Value | HR (95% CI) | *P* Value |
| | *Adjusted for age, sex, and number of beats* | | | | | | | | |
| PVC Count | 1 | 0.9 (0.8-1.1) | 0.584 | 1.4 (1.1-1.8) | 0.005 | 1.6 (1.1-2.3) | 0.014 | 1.3 (1.0-1.8) | 0.083 |
| | 2-4 | 1.1 (0.9-1.3) | 0.456 | 1.4 (1.1-1.8) | 0.01 | 1.4 (0.9-2.2) | 0.118 | 1.7 (1.2-2.4) | 0.002 |
| | 5-10 | 1.2 (0.9-1.6) | 0.281 | 1.9 (1.3-2.6) | <0.001 | 1.9 (1.1-3.4) | 0.018 | 1.6 (1.0-2.6) | 0.039 |
| | 11-25 | 1.1 (0.7-1.6) | 0.684 | 2.1 (1.4-3.1) | <0.001 | 2.9 (1.6-5.1) | <0.001 | 2.2 (1.3-3.8) | 0.004 |
| | >25 | 1.2 (0.8-1.7) | 0.317 | 3.2 (2.3-4.4) | <0.001 | 3.7 (2.3-6.2) | <0.001 | 2.7 (1.7-4.3) | <0.001 |
| | *Adjusted for clinical and ECG exercise test variables, and number of beats* | | | | | | | | |
| PVC Count | 1 | 1.0 (0.8-1.2) | 0.683 | 1.3 (1.1-1.7) | 0.014 | 1.5 (1.1-2.2) | 0.022 | 1.3 (0.9-1.8) | 0.135 |
| | 2-4 | 1.1 (0.9-1.4) | 0.385 | 1.3 (1.0-1.7) | 0.061 | 1.3 (0.9-2.1) | 0.185 | 1.6 (1.1-2.2) | 0.009 |
| | 5-10 | 1,2 (0.9-1.6) | 0.240 | 1.8 (1.3-2.5) | <0.001 | 1.9 (1.1-3.2) | 0.028 | 1.6 (1.0-2.5) | 0.073 |
| | 11-25 | 1.1 (0.7-1.6) | 0.657 | 1.8 (1.2-2.7) | 0.003 | 2.7 (1.5-4.8) | <0.001 | 2.0 (1.2-3.4) | 0.013 |
| | >25 | 1.2 (0.8-1.7) | 0.337 | 2.6 (1.9-3.6) | <0.001 | 3.4 (2.0-5.6) | <0.001 | 2.4 (1.5-3.8) | <0.001 |

There were 1297 (2.7%) myocardial infarction, 276 (0.6%) life-threatening ventricular arrhythmia, 691 (1.4%) heart failure, and 385 (0.8%) cardiovascular death events during follow-up.
Number of beats corresponds to total number of heartbeats (normal + PVCs) recorded during the exercise phase of the stress test. HR: Hazard Ratio, CI: Confidence Interval

Supplemental Table 6: Adjusted Hazard Ratios of Myocardial Infarction, Heart Failure, and Life threatening Ventricular Arrhythmia to PVC count during recovery

| | | Myocardial Infarction | | Heart Failure | | Life threatening Ventricular Arrhythmia | | Cardiovascular Death | |
|---|---|---|---|---|---|---|---|---|---|
| | | HR (95% CI) | *P* Value | HR (95% CI) | *P* Value | HR (95% CI) | *P* Value | HR (95% CI) | *P* Value |
| *Adjusted for age, sex, and number of beats* | | | | | | | | | |
| PVC Count | 1 | 1.1 (0.8-1.3) | 0.614 | 1.6 (1.2-2.1) | <0.001 | 1.1 (0.7-1.8) | 0.757 | 1.4 (1.0-2.1) | 0.047 |
| | 2-4 | 0.9 (0.7-1.3) | 0.598 | 2.2 (1.6-2.9) | <0.001 | 1.4 (0.8-2.5) | 0.23 | 1.5 (0.9-2.3) | 0.093 |
| | ≥5 | 0.9 (0.6-1.3) | 0.512 | 2.5 (1.8-3.5) | <0.001 | 2.5 (1.5-4.2) | <0.001 | 2.1 (1.3-3.3) | 0.003 |
| *Adjusted for clinical and exercise test variables, and number of beats* | | | | | | | | | |
| PVC Count | 1 | 1.1 (0.9-1.4) | 0.475 | 1.5 (1.1-1.9) | 0.003 | 1.0 (0.6-1.7) | 0.885 | 1.3 (0.9-1.9) | 0.109 |
| | 2-4 | 0.9 (0.7-1.3) | 0.617 | 2.0 (1.5-2.7) | <0.001 | 1.3 (0.8-2.3) | 0.324 | 1.4 (0.9-2.2) | 0.183 |
| | ≥5 | 0.9 (0.6-1.4) | 0.650 | 2.3 (1.6-3.1) | <0.001 | 2.2 (1.3-3.8) | 0.003 | 1.9 (1.1-3.1) | 0.014 |

There were 1297 (2.7%) myocardial infarction, 276 (0.6%) life-threatening ventricular arrhythmia, 691 (1.4%) heart failure, and 385 (0.8%) cardiovascular death events during follow-up. Number of beats corresponds to total number of heartbeats (normal + PVCs) recorded during the recovery phase of the stress test. HR: Hazard Ratio, CI: Confidence Interval

Supplemental Table 7: Adjusted Hazard Ratios for Cardiovascular Death to high-grade PVCs during exercise and recovery.

| | Cardiovascular Death | |
|---|---|---|
| | **HR (95% CI)** | ***P* Value** |
| *Adjusted for age, sex, and number of beats* | | |
| High-grade PVCs during exercise | 2.1 (1.5-3.0) | <0.001 |
| High-grade PVCs during recovery | 2.1 (1.3-3.4) | 0.003 |
| *Adjusted for clinical and exercise test variables, and number of beats* | | |
| High-grade PVCs during exercise | 1.9 (1.3-2.7) | <0.001 |
| High-grade PVCs during recovery | 1.9 (1.2-3.2) | 0.016 |

There were 385 (0.8%) cardiovascular deaths during follow-up. HR: Hazard Ratio, CI: Confidence Interval

Supplemental Table 8: Sensitivity analyses: effect of high PVC burden at rest on association strength of exericse PVCs for MACE and all-cause mortality.

| | | Original model | | Original model + PVC burden at rest | |
|---|---|---|---|---|---|
| | | HR (95% CI) | *P* Value | HR (95% CI) | *P* Value |
| | *MACE* | | | | |
| PVC Count | 1 | 1.1 (1.0-1.3) | 0.112 | 1.1 (1.0-1.3) | 0.121 |
| | 2-4 | 1.2 (1.0-1.4) | 0.022 | 1.2 (1.0-1.4) | 0.026 |
| | 5-10 | 1.4 (1.1-1.7) | 0.004 | 1.4 (1.1-1.7) | 0.007 |
| | 11-25 | 1.5 (1.1-1.9) | 0.006 | 1.4 (1.1-1.9) | 0.012 |
| | >25 | 1.8 (1.4-2.3) | <0.001 | 1.7 (1.3-2.2) | <0.001 |
| | *All-cause mortality* | | | | |
| PVC Count | 1 | 1.0 (0.9-1.2) | 0.986 | 1.0 (0.9-1.2) | 0.989 |
| | 2-4 | 1.1 (0.9-1.3) | 0.188 | 1.1 (0.9-1.3) | 0.192 |
| | 5-10 | 1.1 (0.8-1.3) | 0.627 | 1.1 (0.8-1.3) | 0.637 |
| | 11-25 | 1.4 (1.1-1.8) | 0.012 | 1.4 (1.1-1.8) | 0.015 |
| | >25 | 1.5 (1.2-1.9) | 0.002 | 1.5 (1.1-1.9) | 0.004 |

MACE: major adverse cardiovascular events. Original model: clinical and ECG stress test factors, and number of beats during exercise; Original model + PVC burden at rest: adjusted for all variables in the original model + a binary variable indicating the presence of high PVC burden at rest. HR: Hazard Ratio, CI: Confidence Interval

Supplemental Table 8: Sensitivity analyses: effect of high PVC burden at rest on association strength of recovery PVCs for MACE and all-cause mortality.

| | | Original model | | Original model + high PVC burden at rest | |
|---|---|---|---|---|---|
| | | **HR (95% CI)** | ***P* Value** | **HR (95% CI)** | ***P* Value** |
| | *MACE* | | | | |
| PVC Coun | 1 | 1.2 (1.0-1.4) | 0.028 | 1.2 (1.0-1.4) | 0.038 |
| | 2-4 | 1.3 (1.0-1.6) | 0.038 | 1.2 (1.0-1.5) | 0.075 |
| | ≥5 | 1.5 (1.2-1.9) | 0.001 | 1.4 (1.1-1.8) | 0.010 |
| | *All-cause mortality* | | | | |
| PVC Coun | 1 | 1.1 (0.9-1.3) | 0.363 | 1.1 (0.9-1.3) | 0.377 |
| | 2-4 | 1.1 (0.9-1.4) | 0.430 | 1.1 (0.9-1.4) | 0.471 |
| | ≥5 | 1.5 (1.2-1.8) | 0.002 | 1.4 (1.1-1.8) | 0.005 |

MACE: major adverse cardiovascular events. Original model: clinical and ECG stress test factors, and number of beats during recovery; Original model + PVC burden at rest: adjusted for all variables in the original model + a binary variable indicating the presence of high PVC burden at rest. HR: Hazard Ratio, CI: Confidence Interval
